# A re-examination of the impact of COVD-19 deaths on the computation of average life expectancy

**DOI:** 10.1101/2022.01.20.22269578

**Authors:** Arni S.R. Srinivasa Rao, Steven G. Krantz, David A. Swanson

## Abstract

It is natural to question the impact of COVID-19 on life expectancy. However, a newborn during the 2020-2021 period need not experience the same level of adult mortality found in 2020-2021 because there may be zero COVID-19 related deaths when the newborn reaches adulthood. Thus, life expectancy lost due to COVID-19 cannot be found simply by incorporating excess deaths due to COVID-19 and re-doing the life table computations because: (1) we know that the COVID-19 deaths need not occur every year for the next 20-25 years; and (2) once an adult, a newborn in 2021/2022 need not experience the same mortality rate that current middle and older aged COVID-19 patients experience. Using U.S. data as an example, we estimate an average of 29.68 years of life was lost to those aged 18-64 who died from COVID-19 in the U.S., noting that 74 % of the reported deaths of 18-64 occurred among 50-64 years and 10 % below 40 years. Instead of computing life expectancy years lost due to COVID-19, we recommend computing life years lost due to COVID-19.

## Introduction

As of January 19^th^, 2022 the total worldwide COVID-19 related deaths from the beginning of the pandemic in 2019 are around 5.55 million, and total infections are around 334 million (Ritchie et al., 2022). We also note that there have been under-reporting and under-diagnosis of these numbers (Krantz and Rao 2020, Finelli and Swerdlow 2023). Some researchers have measured the impact of these excess deaths on population life expectancy, while others argued that life expectancy is reduced due to COVID-19 deaths (Trias-Llimós and Bilal 2020, Andrasfay and Goldman 2021, Castro et al., 2021, Venkataramani, O’Brien and Tsai 2021). In general, the approach considered in these studies is one of the commonly accepted techniques using period life tables (Chan, Cheng, and Martin 2021, Yusuf, Martins, and Swanson 2014). However, the middle and older-aged excess deaths during 2020-21 influence the computation of life expectancy of newborn babies of 2020-21 in all of them. In essence, these studies assume that the current mortality rates among adults will have the same pattern when a current newborn reaches adulthood, and, accordingly, survival probabilities were computed. However, a newborn in 2020 need not be influenced by COVID-19 excess adult deaths during 2020-21 because these deaths may not occur every year for the next 20-30 years. Misra (1998) argued that one needs to consider the lives lost due to a cause of interest and then compute the mean life lost by all deaths.

In general, we agree with this approach, and extend it by proposing that each individual life lost due to COVID-19 should be associated with computing the impact of excess deaths due to COVID-19. As an example, we consider the distribution of deaths in the U.S. until November, 2021 and construct the life years lost due to COVID-19 using the person-years data and adjusting the average life years lived relative to the corresponding COVID-19 deaths. Figure 1 provides a description of the data and our idea of computing average person-years lost due to COVID-19. Computational details are found in the Appendix.

**Figure 1.**
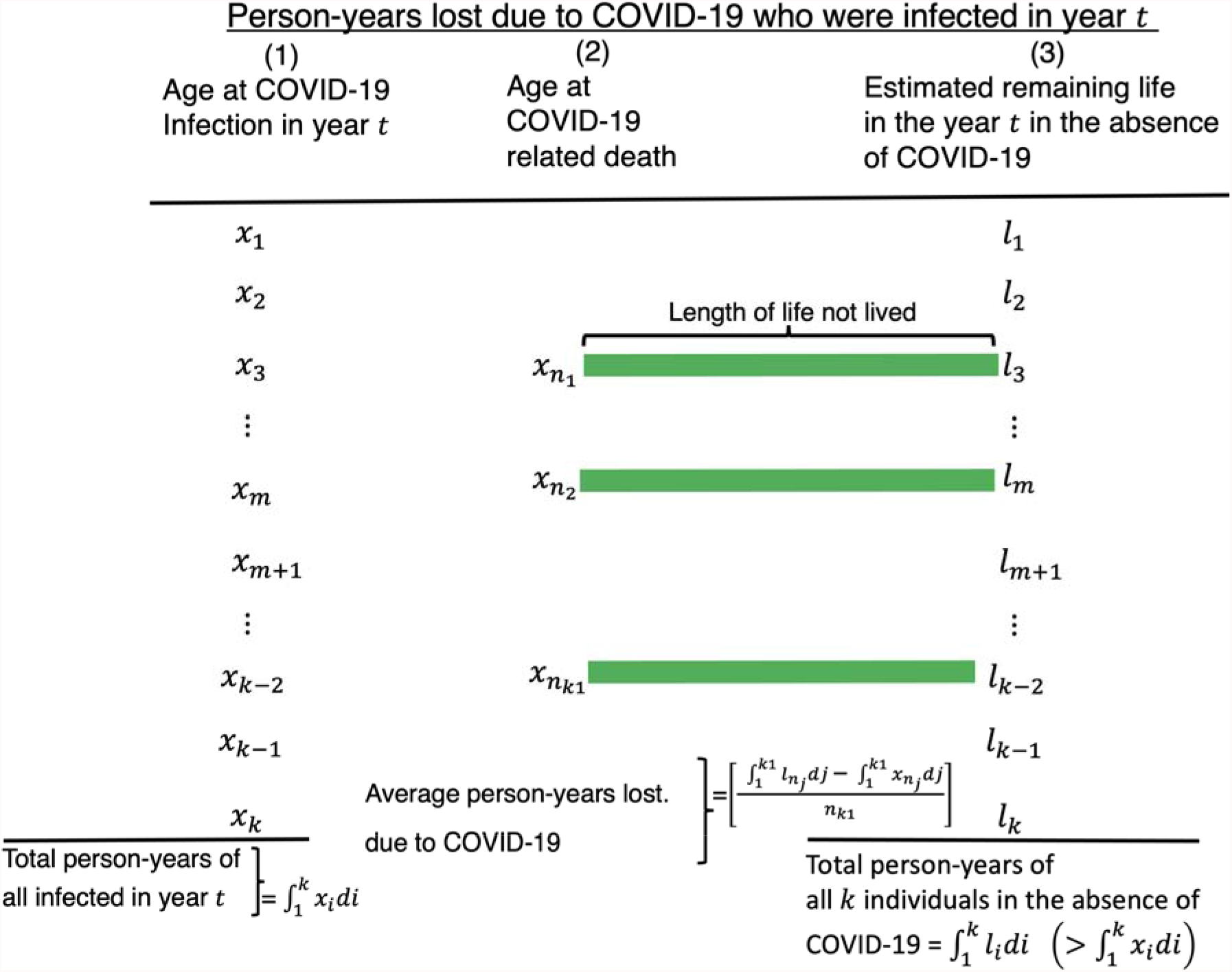
Computation of person-years lost by COVID-19 deaths. The details of computations are explained in Materials and Methods.

Following Goldstein and Lee (2020) and Heuveline (2021), we next compute the loss of expected remaining life due to COVID-19.

### Average life lost due to COVID-19

Computation of remaining loss of life due to COVID-19 involves some level of uncertainty. However, this approach provides a better measure than simply computing the loss of years in life expectancy using a contemporary period life table. The level of uncertainty in computing this loss function is unavoidable because the estimation of remaining-life years requires the computations of future survival probabilities, which are usually obtained through life table approaches.

Consider the 764,715 deaths among those 18 years and over due to COVID-19 in the U.S. until November 17, 2021 (Statistica 2022), for which the age at death is displayed in Table 1.

**Table 1.**
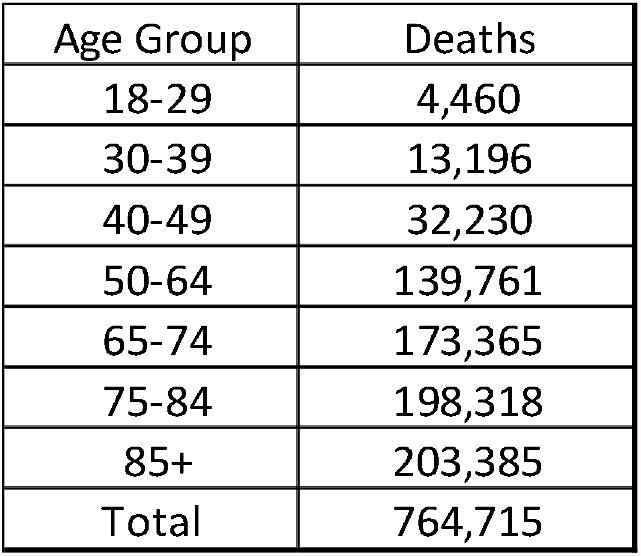
U.S. COVID-19 Deaths by Age to 17 November 2021

Using the National Vital Statistics reports released in November 2020 (Arias and Xu 2020), we find that the U.S. population aged 18 years and over had 6,086,463 person-years remaining to live in the absence of COVID-19.

Using standard life table methods (Yusuf, Martins, and Swanson 2014), we computed the person-years of life lost (PYLL) by age group to the decedents shown in Table 1. They are: 250,599 PYLL to those aged 18-29; 605,323 to those aged 30-39; 1,181,695 to those aged 40-49; 3,612,596 to those aged 50-64; 1,868,922 to those aged 65-74; 1,902,847 to those aged 75-84; and 812,586 PYLL to those decedents aged 85+.

The weighted average number of years lost due to those who are approximately 41.5 years (the mid-point of the age range 18-65) is 29.68 years. This age-group is important because it corresponds closely to the definition of the “working age” population, which is defined as the age group, 15-64 (Organization for Economic Cooperation and Development, 2022) and used in the U.S and other countries as the basis for many tax and economic policies. U.S. life expectancy (all sexes) is 76 years in the absence of COVID-1 (Arias and Xu 2020). So, on average, a person aged 41.5 can expect to live 34.5 more years in the absence of COVID-19. Our computations show, on average, that 29.68 years of these 34.5 years were lost due to COVID-19. Most of these deaths (74%) occurred in age group 50-64 while only about 10 % of these deaths occurred among people below 40 years of age.

The average remaining years that would have been there for the individuals in the U.S. who died due to COVID-19 in various age groups are as follows: 56.2 (aged 18-29); 45.9 (aged 30-39); 36.9 (aged 40-49); 25.8 (aged 50-64); 10.8 (aged 65-74); 9.6 (aged 75-84); and 4.0 (aged 85+).

## Discussion and Limitations

In this paper, we provide an estimate of a loss of average life due to excess deaths due to COVID-19 in the U.S. that represents an improvement over the period life table-based methods that measure impact on life expectancies. However, it has limitations that are largely unavoidable. One such limitation includes assuming that the age at death of individuals within an age group is uniformly distributed. That means the remaining lifetime probability for an individual aged 39.2 years is the same as the remaining probability for an individual aged 39.9. The other standard limitations that apply to the period life table also are assumed to hold (Arias and Xu 2020, Yusuf Martins and Swanson 2014).

## Data Availability

All data produced in the present work are contained in the manuscript and generated from cited sources available to anyone.

https://www.cdc.gov/nchs/data/nvsr/nvsr69/nvsr69-12-508.pdf

https://www.statista.com/statistics/1191568/reported-deaths-from-covid-by-age-us/

https://ourworldindata.org/explorers/coronavirus-data-explorer

## Acknowledgments

Rao and Krantz thank the U.S. National Science Foundation for funding through the project DMS-2140493.

## Appendix

The computation of average life lost due to COVID-19 deaths during 2020-2021 proceeds as follows:

Suppose *x*_*i*_ for *i* = 1,2,…, *k*, be the *i*^*th*^ individual infected with COVID-19 in year *t*. Let X = {*x*_*i*_ : *i* = 1,2,…, *k*}. Suppose there are *k*_1_ deaths among the set of individuals in *A*. Let 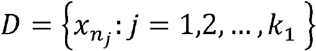, where 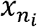 is the age at death of *j*^*th*^ individual. Note that,

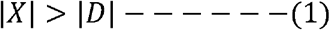

where |*x*| and |*D*| are the sizes of the sets *x* and *D*, respectively. Let *l*_*i*_ be the remaining years to live for the *i*^*th*^ individual in the absence of COVID-19 and computed for the year *t*. Three quantities of person-years that we compute are as follows:

The total person-years lived by the individuals in *X* is

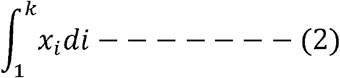

The total person-years to be lived by the individuals in *X* is

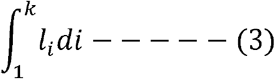

The average person-years lost due to the set *D* is computed as

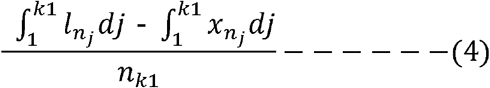

The quantity 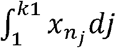 in (4) is computed from the actual deaths, and the quantity 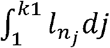 in (4) is computed from a complete life table computed for year *t* in the absence of COVID-19 deaths. There are a few more assumptions involved in computing this integral. The probability of occurrence of an event *D* is conditional to the occurrence of the event *X*, we compute the conditional probability, *P*(*D*/*X*) as

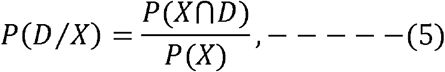

where the event of both *X* and *D* occurring is given by *X*∩*D*.

Since *X*∩*D* = *D*, we can write (5) as

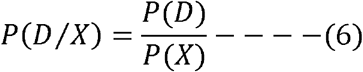

In (6), the quantity *P*(*D*) can be approximated as

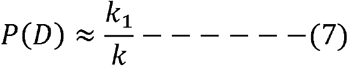

and

the quantity *P*(*X*) can be approximated as

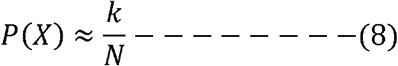

where the size of mid-year population in the year *t* is *N*.

## References

Andrasfay T., and N. Goldman (2021). Association of the COVID-19 Pandemic with Estimated Life Expectancy by Race/Ethnicity in the United States. JAMA Netw Open. 4(6):e2114520. doi:10.1001/jamanetworkopen.2021.14520

Angulo F., L. Finelli, and D. Swerdlow (2021). Estimation of US SARS-CoV-2 Infections, Symptomatic Infections, Hospitalizations, and Deaths Using Seroprevalence Surveys. JAMA Netw Open 4(1):e2033706. doi:10.1001/jamanetworkopen.2020.33706

Arias, E., and J. Xu. (2020). U.S. Life Tables, 2018. National Vital Statistics Reports 69 (12). Centers for Disease Control. https://www.cdc.gov/nchs/data/nvsr/nvsr69/nvsr69-12-508.pdf

Castro, M., S. Gurzenda, C. Turra, T. Andrasfay, and N. Goldman (2021). Reduction in life expectancy in Brazil after COVID-19. Nature Medicine 27, 1629–1635 https://doi.org/10.1038/s41591-021-01437-z

Chan E., D. Cheng, and J. Martin. (2021) Impact of COVID-19 on excess mortality, life expectancy, and years of life lost in the United States. PLoS ONE 16(9): e0256835. https://doi.org/10.1371/journal.pone.0256835.

Goldstein J., and R. Lee (2020. Demographic perspectives on the mortality of COVID-19 and other epidemics. Proceedings of the National Academy of Sciences of the United States of America.117(36): 22035–22041. pmid:32820077.

Heuveline P. (2021) The Mean Unfulfilled Lifespan (MUL): A new indicator of the impact of mortality shocks on the individual lifespan, with application to mortality reversals induced by COVID-19. PLoS ONE 16(7): e0254925. https://doi.org/10.1371/journal.pone.0254925.

Krantz, S.G, Rao, A.S.R.S. (2020). Level of underreporting including underdiagnosis before the first peak of COVID-19 in various countries: Preliminary retrospective results based on wavelets and deterministic modeling. Infection Control & Hospital Epidemiology, 41(7), 857–859. doi:10.1017/ice.2020.116

Misra, B.D. (1998). An Introduction to the Study of Population. South Asian Publishers, New Delhi, 1995.

Organization for Economic Cooperation and Development. (2022), Working age population (indicator). doi: 10.1787/d339918b-en (Accessed on 19 January 2022)

Pifarréi Arolas, H. E. Acosta, G. López-Casasnovas, A. Lo, C. Nicodemo, T. Riffe and M. Myrskylä. (2021). Years of life lost to COVID-19 in 81 countries. Scientific Reports 11: 3504 https://doi.org/10.1038/s41598-021-83040-3.

Ritchie, H., E. Mathieu, L. Rodés-Guirao, C. Appel, C. Giattino, E, Ortiz-Ospina, J. Hasell, B. Macdonald, D. Beltekian and M. Roser. (2022). Coronavirus Pandemic (COVID-19). Coronavirus Pandemic (COVID-19) – the data -Statistics and Research - Our World in Data (accessed January 19th, 2022).

Statistica (2022). https://www.statista.com/statistics/1191568/reported-deaths-from-covid-by-age-us/.

Trias-Llimós S, and U. Bilal. (2020). Impact of the COVID-19 pandemic on life expectancy in Madrid (Spain), Journal of Public Health 42 (3): 635–636, https://doi.org/10.1093/pubmed/fdaa087.

Venkataramani A., R. O’Brien, A. Tsai (2021). Declining Life Expectancy in the United States: The Need for Social Policy as Health Policy. JAMA 325 (7):621– 622. doi:10.1001/jama.2020.26339.

Yusuf, F, J. Martins, and D. Swanson. (2014). Multiple Decrement Life Tables, pp. 215–299 in Methods of Demographic Analysis. Springer B.V. Press. Dordrecht, Heidelberg, London, and New York.

